# Knowledge, attitudes and practices associated with the COVID-19 among slum dwellers resided in Dhaka City: A Bangladeshi interview-based survey

**DOI:** 10.1101/2020.09.15.20195255

**Authors:** Md. Saiful Islam, Md. Galib Ishraq Emran, Md. Estiar Rahman, Rajon Banik, Md. Tajuddin Sikder, Lee Smith, Sahadat Hossain

## Abstract

**Background:** The emergent COVID-19 has impacted unprecedentedly to all classes of people. Slum-dwellers’ knowledge, attitudes and practices (KAP) towards COVID-19 are currently poorly understood. To investigate the KAP towards COVID-19 among slum dwellers resided in Dhaka City, Bangladesh.

**Methods:** A cross-sectional offline survey was carried out enrolling 406 slum dwellers (53.2% male; mean age=44.9 years [SD=12.1]; age range=18-85 years) between August and September, 2020. The face to face interview was conducted to collect data from 6 selected slum areas in Dhaka City using convenience sampling. The questionnaire consisted of informed consent along with questions concerning observational checklists, socio-demographics, and KAP.

**Results:** A sizeable minority were observed without wearing face masks during the survey periods (18.2%) and a vast portion (97.5%) without any hand protection. The mean scores of KAP were 6.1±2.6 (out of 17), 12.3±1.7 (out of 14), and 9.8±1.6 (out of 12), respectively. Moreover, the KAP were strongly and positively correlated with each other.

**Conclusions:** The findings revealed that the majority of slum dwellers in Bangladesh have limited knowledge of COVID-19. Poor practices (i.e., face mask and hand protection) were directly observed during the survey. The findings suggest the immediate implementation of health education programs and adequate interventions.

## Introduction

Coronavirus disease 2019 (COVID-19) outbreak, caused by the new coronavirus strain SARS-CoV-2, has become a serious public health concern worldwide (1). The outbreak was first revealed in Wuhan city, in the Hubei Province of China, in late December 2019. The outbreak soon spread to the whole country, reached beyond the border, and eventually, the World Health Organization (WHO) declared the outbreak a pandemic on March 11, 2020 (2). At the time of writing (September 20, 2020), this pandemic has affected 216 countries, areas, and territories with over 28.9 million confirmed cases and 922,252 deaths recorded globally (3). On March 8, 2020, Bangladesh announced the first case of COVID-19 (4–6). Since then, the numbers of new cases are rising in the country. As of September 20, 2020, the country has recorded 339,332 positive cases of COVID-19 and 4,759 deaths domestically (7).

As yet, no vaccine or specific treatment is available for the control and management of the disease (8). The most effective measure of controlling the spread of the virus is to protect oneself from being exposed to COVID-19. People’s knowledge, attitudes, perception and practices regarding the disease are the keys to ensuring success in the battle against the deadly disease. The WHO has prescribed some general guidelines for all sections of people to remain protected from the disease.

Most affected countries around the world have adopted non-therapeutic measures including lockdown, social distancing, self-isolation, or quarantine, in order to combat the spread of COVID-19 (9). The Bangladeshi government declared a nationwide lockdown from March 26 to May 30, 2020, extending it seven times (4,10,11). Furthermore, the government has extended restrictions imposed on public activities and movement across the country until August 31, 2020, to limit the spread of COVID-19 (12).

A study investigating Chinese people’s knowledge, attitude, and practice regarding COVID-19 concealed that public attitudes to obedience to government measures to combat the epidemic were significantly influenced by the level of knowledge about COVID-19 (13). More knowledge was also found to be correlated with more optimistic attitudes towards COVID-19 preventive practices (13,14). In Bangladesh, a recent study showed that a large proportion of people had limited knowledge of COVID-19 transmission and onset of symptoms and had positive perceptions of COVID-19 (15). Another Bangladeshi study found despite 54.9% respondents kept good knowledge but the attitude and practices were not impressive mainly because of poor knowledge, non-scientific, and orthodox religious belief (16).

Dhaka (where the present study carried out), the capital city of Bangladesh, has more than 3,000 slums inhabited by over 6 lakh people (17). The slums are densely populated; around 75 percent of households live in one room (18). Apart from this, shared kitchens, toilets, communal water sources, lack of education and economic vulnerabilities make the slum residents the most vulnerable to COVID-19. As mentioned above, people’s knowledge, attitudes, perception and practices are crucial to prevent the novel coronavirus, it is therefore important to investigate the slum residents’ knowledge, attitudes and practices associated with COVID-19.

To date, there is no conducted among slum dwellers in Bangladesh investigating their knowledge, attitudes and practices towards COVID-19. Consequently, the present study aimed to assess knowledge, attitudes and practices associated with COVID-19 among slum dwellers in Bangladesh.

## Methods

### Participants and procedures

A cross-sectional offline survey was carried out enrolling 406 slum dwellers between August and September, 2020. A convenience sampling technique was employed to draw the sample from 6 selected slum areas (i.e., Khurshid Bari Bosti, Shorgochera Bosti, Balur Maath Songlongno Bosti, Aziz Shaheber Bosti Bari, Pinur Bosti, and Fighter Bosti) located in Dhaka city, Bangladesh. The face to face interview was conducted to gather information from participants with maintaining proper precaution and spatial distancing using a semi-structured questionnaire including informed consent. The inclusion criteria of the participants were that individuals had to be aged ≥18 years, and be slum dwellers. Exclusion criteria included individuals being <18 years old and not consenting the survey willingly. After obtaining informed consent, 410 participants were interviewed. After removing incomplete surveys, data from 406 participants (53.2% male; age range 18-85 years; mean age 44.9 years [SD=12.1]) were included in analyses.

### Measures

The questionnaire containing informed consent along with three sections (i.e., Observational checklists, Socio-demographic information, KAP) was utilized to collect data during the interview periods.

### Observational checklists

The observational checklists were espied during the survey periods, consisted of the two items ‘yes/no’ questions regarding participants’ practices in terms of wearing face mask and hand protection (e.g., using globs, hand sanitizer, etc.).

### Socio-demographic information

Some questions related to socio-demographics were asked during the survey including gender (male/female), age (later categorized: 18-40 years and >40 years), education (no formal education, primary level [1-5 grades], and secondary level [6-10 grades]), occupation, marital status (unmarried/married/divorced or widows or widowers), family type (nuclear/joint), and monthly family income (later categorized: <5,000 Bangladeshi Taka [BDT], 6,000-15,000 BDT and >15,000 BDT).

### Knowledge, attitudes, and practices (KAP)

Participants’ knowledge, attitudes, and practices towards the COVID-19 were measured using a total of 28 items structured questions (including 17-item for knowledge, 7-item for attitudes, and 6-item for practices) based on the two prior studies (19,20) conducted in Bangladesh as well as the infection prevention and control measures for COVID-19 by World Health Organization (21). A pilot study was conducted before the inauguration of the final data collection.

The knowledge section comprised of 17-item ‘true/false’ questions concerning mode of transmission (5-item), clinical manifestations (7-item), incubation period (1-item), and risk factors (3-items) (e.g., “*COVID-19 can spread through respiratory droplets of infected individuals*.”, see details in Table 2). The correct answer (’*True*’) was coded as 1, while the wrong answer (’*False*’) was coded as 0 (19). The total score ranges from 0-17, with an overall greater score indicates more favorable knowledge towards the COVID-19.

**Table 1.** Distribution of all examined variables (N=406)

| **Characteristics** | **n** | **(%)** |
| --- | --- | --- |
| **Gender** |  |  |
| Male | 216 | (53.2) |
| Female | 190 | (46.8) |
| **Age** |  |  |
| 18-40 years | 182 | (44.8) |
| >40 years | 224 | (55.2) |
| **Education** |  |  |
| No formal education | 53 | (13.1) |
| Primary level (1-5 grades) | 301 | (74.1) |
| Secondary level (6-10 grades) | 52 | (12.8) |
| **Occupation** |  |  |
| Housewife | 53 | (13.1) |
| Workers | 92 | (22.7) |
| Day laborer | 27 | (6.7) |
| Small shop keeping | 104 | (25.6) |
| Rickshaw Puller | 107 | (26.4) |
| Others | 23 | (5.7) |
| **Marital status** |  |  |
| Unmarried | 20 | (4.9) |
| Married | 352 | (86.7) |
| Divorced | 34 | (8.4) |
| **Family type** |  |  |
| Nuclear | 361 | (88.9) |
| Joint | 45 | (11.1) |
| **Monthly family income** |  |  |
| <5,000 BDT | 107 | (26.4) |
| 6,000-15,000 BDT | 250 | (61.6) |
| >15,000 BDT | 49 | (12.1) |
| **Using mask** |  |  |
| Yes | 332 | (81.8) |
| No | 74 | (18.2) |
| **Hand protection** |  |  |
| Yes | 10 | (2.5) |
| No | 396 | (97.5) |
| **Knowledge (Mean±SD)** |  | (6.1±2.6) |
| **Attitude (Mean±SD)** |  | (12.3±1.7) |
| **Practice (Mean±SD)** |  | (9.8±1.6) |

Note: BDT=Bangladeshi Taka

**Table 2.** Knowledge and gender difference of participants (N=406)

| Characteristics | Total N=406 | Male | Female | $\chi^2$ | df | p-value |
| --- | --- | --- | --- | --- | --- | --- |
|  | n (%) | n (%) | n (%) |  |  |  |
| COVID-19 can spread through respiratory droplets of infected individuals. |  |  |  |  |  |  |
| True | 336 (82.8) | 178 (82.4) | 158 (83.2) | 0.040 | 1 | 0.842 |
| False | 70 (17.2) | 38 (17.6) | 32 (16.8) |  |  |  |
| COVID-19 can spread through close contact of infected persons |  |  |  |  |  |  |
| True | 69 (17.0) | 54 (25.0) | 15 (7.9) | 20.966 | 1 | <0.001 |
| False | 337 (83.0) | 162 (75.0) | 175 (92.1) |  |  |  |
| COVID-19 can spread via infected surfaces |  |  |  |  |  |  |
| True | 4 (1.0) | 3 (1.4) | 1 (.5) | 0.771 | 1 | 0.626* |
| False | 402 (99.0) | 213 (98.6) | 189 (99.5) |  |  |  |
| COVID-19 can spread through touching face, mouth, and eyes by infected hands |  |  |  |  |  |  |
| True | 20 (4.9) | 17 (7.9) | 3 (1.6) | 8.543 | 1 | 0.003 |
| False | 386 (95.1) | 199 (92.1) | 187 (98.4) |  |  |  |
| COVID-19 can spread through poor maintaining of spatial distancing |  |  |  |  |  |  |
| True | 9 (2.2) | 8 (3.7) | 1 (.5) | 4.708 | 1 | 0.04* |
| False | 397 (97.8) | 208 (96.3) | 189 (99.5) |  |  |  |
| Difficulty breathing or shortness of breath is a serious symptom of COVID-19 |  |  |  |  |  |  |
| True | 180 (44.3) | 110 (50.9) | 70 (36.8) | 8.124 | 1 | 0.004 |
| False | 226 (55.7) | 106 (49.1) | 120 (63.2) |  |  |  |
| Chest pain is a serious symptom of COVID-19 |  |  |  |  |  |  |
| True | 54 (13.3) | 38 (17.6) | 16 (8.4) | 7.374 | 1 | 0.007 |
| False | 352 (86.7) | 178 (82.4) | 174 (91.6) |  |  |  |
| Fever is a common symptom of COVID-19 |  |  |  |  |  |  |
| True | 364 (89.7) | 197 (91.2) | 167 (87.9) | 1.193 | 1 | 0.275 |
| False | 42 (10.3) | 19 (8.8) | 23 (12.1) |  |  |  |
| Dry cough is a common symptom of COVID-19 |  |  |  |  |  |  |
| True | 312 (76.8) | 180 (83.3) | 132 (69.5) | 10.913 | 1 | 0.001 |
| False | 94 (23.2) | 36 (16.7) | 58 (30.5) |  |  |  |
| Fatigue/tiredness is a common symptom of COVID-19 |  |  |  |  |  |  |
| True | 44 (10.8) | 35 (16.2) | 9 (4.7) | 13.755 | 1 | <0.001 |
| False | 362 (89.2) | 181 (83.8) | 181 (95.3) |  |  |  |
| Aches and pains are less common symptom of COVID-19 |  |  |  |  |  |  |
| True | 42 (10.3) | 29 (13.4) | 13 (6.8) | 4.724 | 1 | 0.030 |
| False | 364 (89.7) | 187 (86.6) | 177 (93.2) |  |  |  |
| Sore throat is a less common symptom of COVID-19 |  |  |  |  |  |  |
| True | 306 (75.4) | 170 (78.7) | 136 (71.6) | 2.764 | 1 | 0.096 |
| False | 100 (24.6) | 46 (21.3) | 54 (28.4) |  |  |  |
| Diarrhea is a less common symptom of COVID-19 |  |  |  |  |  |  |
| True | 28 (6.9) | 22 (10.2) | 6 (3.2) | 7.774 | 1 | 0.005 |
| False | 378 (93.1) | 194 (89.8) | 184 (96.8) |  |  |  |
| Symptoms appear after 2-14 days |  |  |  |  |  |  |
| True | 315 (77.6) | 175 (81.0) | 140 (73.7) | 3.127 | 1 | 0.077 |
| False | 91 (22.4) | 41 (19.0) | 50 (26.3) |  |  |  |
| Older persons are at risk for developing COVID-19 |  |  |  |  |  |  |
| True | 379 (93.3) | 207 (95.8) | 172 (90.5) | 4.586 | 1 | 0.032 |
| False | 27 (6.7) | 9 (4.2) | 18 (9.5) |  |  |  |
| Pregnant women are at risk for developing COVID-19 |  |  |  |  |  |  |
| True | 8 (2.0) | 5 (2.3) | 3 (1.6) | 0.283 | 1 | 0.728 |
| False | 398 (98.0) | 211 (97.7) | 187 (98.4) |  |  |  |
| Individuals with chronic diseases are at risk for developing COVID-19 |  |  |  |  |  |  |
| True | 1 (0.2) | 1 (.5) | 0 (.0) | 0.882 | 1 | 1.00 |
| False | 405 (99.8) | 215 (99.5) | 190 (100.0) |  |  |  |

Note: Fisher's exact test

The attitudes section consisted of 7-item questions regarding the positive attitudes towards the severity and prevention of COVID-19 (e.g., “*Coronavirus is very serious*.” see details in Table 6) with a three-point Likert scale including 0 (“*Disagree*”), 1 (“*Undecided*”), 2 (“*Agree*”) (20), and yielding total scores ranging from 0 to 14. An overall greater score indicates more positive attitudes towards the COVID-19.

The practices section included 6-item questions concerning the preventive measures of COVID-19 (e.g., “*Do you use tissues or hanker chips during coughing/sneezing?*” see details in Table 8) with a three-point Likert scale ranging from 0 (“*Never*”) to 3 (“*Always*”) (20). Practice items’ total score ranges from 0-12, with an overall greater score indicates more frequent practices towards the COVID-19.

### Statistical analyses

All data were coded and analyzed using two statistical software packages (Microsoft Excel 2019, and IBM SPSS Statistics version 25). Microsoft Excel was used to perform data cleaning, coding, editing, and sorting. An Excel file including all variables was imported in SPSS software. Descriptive statistics (e.g., frequencies, percentages, means, standard deviations, etc.) and some first-order analyses (e.g., Chi-square tests, Fisher’s exact tests, etc.) were performed using SPSS software. In addition, *t*-tests or one-way ANOVA tests were performed to determine significant relations of the mean KAP scores with all examined variables. Finally, demographic, and observational checklists’ variables that significantly differed in terms of knowledge, attitudes, and practices scores, were included into hierarchical regression analysis with knowledge, attitudes, and practices, respectively as the dependent variable. Furthermore, a bivariate Pearson correlation was performed along with the total scores of knowledge, attitudes, and practices to investigate the significant relationships with each other.

### Ethics

All procedures of the present study were carried out in accordance with the principle for human investigations (i.e., Helsinki Declaration). Furthermore, the study was conducted in accordance with the ethical guidelines of the Institutional research ethics committee. Formal ethics approval was granted by the Institutional ethics review board of Jahangirnagar University (Ref. No: BBEC, JU/M 2020/COVID-19/(8)5). Participants were informed about the procedure and purpose of the study and the confidentiality of information provided. All participants consented willingly to be a part of the study during the interview periods. All data were collected anonymously and analyzed using the coding system.

## Results

### General characteristics of participants

A total of 406 participants included in the final analysis. Of them, 53.2% were male with their mean age 44.9 years (SD=12.1) ranging from 18-85 years and most had primary level of education (1-5 grades) (74.1%) (Table 1). The majority of participants were rickshaw pullers (26.4%) and had their monthly family income 6,000-15,000 BDT (61.6%). Most were married (86.7%) and lived together with nuclear families (88.9%). A sizeable minority were observed without wearing face masks during the survey periods (18.2%). A vast portion of participants (97.5%) were observed in the survey periods without any hand protection (e.g., hand gloves, hand sanitizer, etc.).

### Knowledge

The mean scores of knowledge were 6.1 (SD=2.6) out of 17 with an overall correct rate of 35.9%. The participants’ source of knowledge regarding the COVID-19 included as follows: mass media (95.6%), friends and neighbors (45.8%), family members (48.8%), internet (2.7%), and social media (1.5%) (see Figure 1). The distribution of each knowledge question along with gender differences was presented in Table 2.

**Figure 1.**
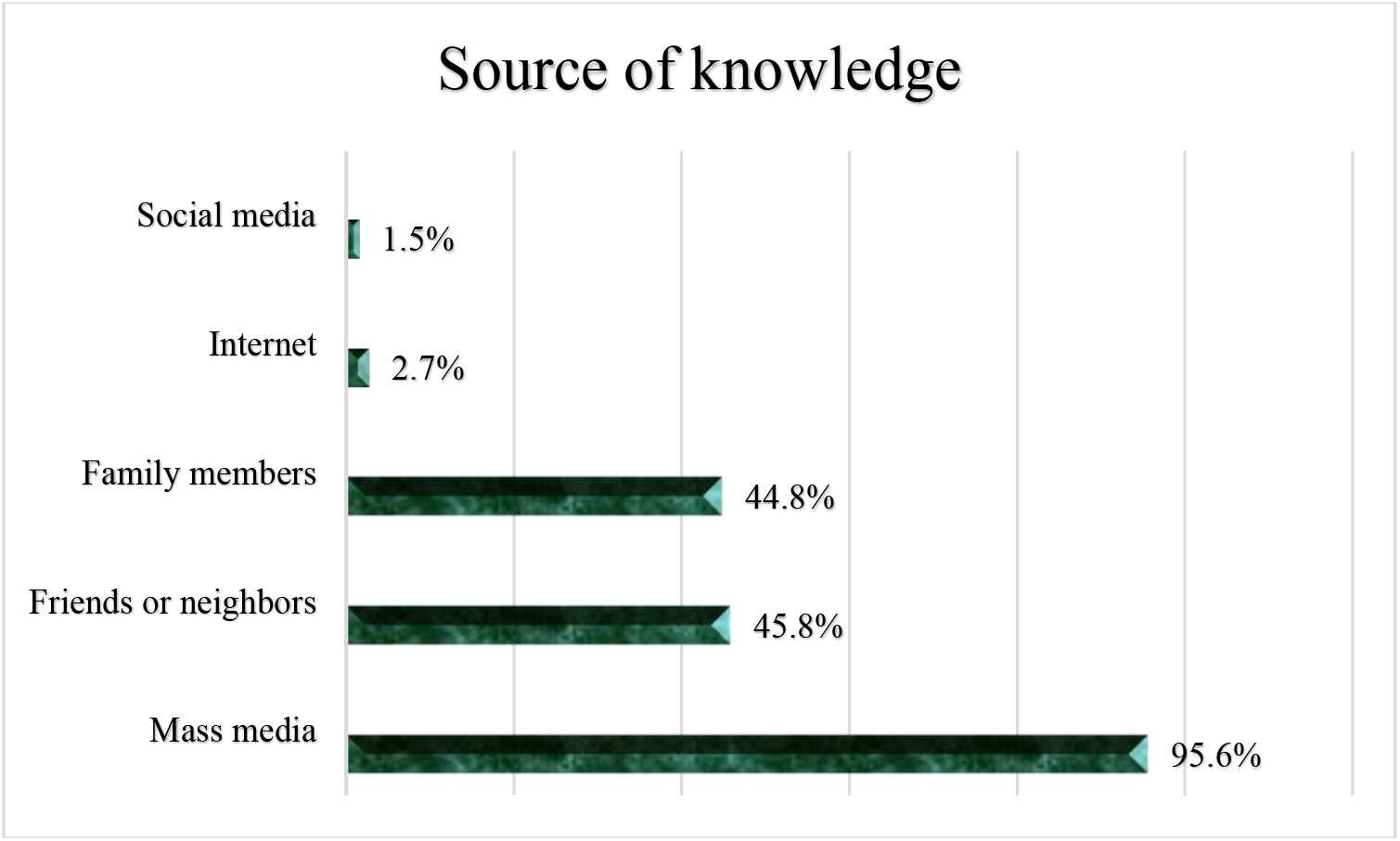
The participants’ source of knowledge regarding the COVID-19.

The participants’ knowledge score was significantly higher among i) males vs. females (6.6±2.6 vs. 5.5±2.3, *p*=0.002), ii) participants with secondary level of education vs. no formal education (6.9±2.8 vs. 5.0±3.5, *p*<0.001), iii) unmarried vs. divorced participants (7.7±2.8 vs. 4.1±2.8, *p*<0.001), and iv) participants those were observed with vs. without the hand protection during the survey periods (7.6±4.1 vs. 6.0±2.5, *p*<0.001) (see Table 3).

**Table 3.**
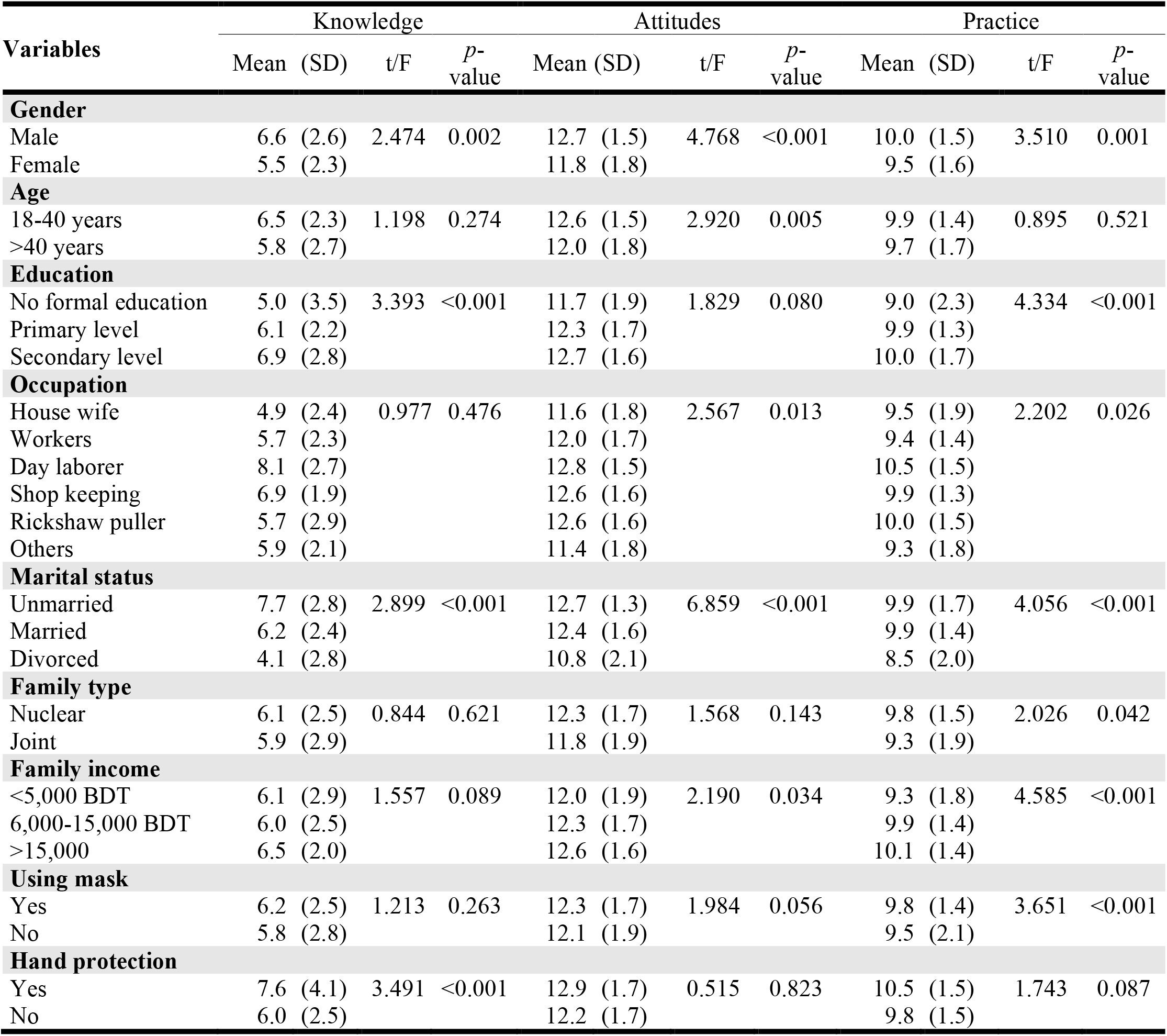
Association between socio-demographic characteristics and mean knowledge, attitudes and practices score (N=406)

The results of hierarchical regression analysis for knowledge are presented in Table 4. Factors that were statistically significant in the group difference analyses (t-tests and ANOVA) were included in a hierarchical regression analysis. Socio-demographic factors (i.e., gender, education, and marital status) were included in Block 1 and Observation checklist (i.e., hand protection) comprised Block 2. The more favorable knowledge was positively associated with males (β=-0.16, *p*<0.01), having secondary level of education (β=0.14, *p*<0.01), and being unmarried (β=-0.19, *p*<0.001). Consequently, hand protection was not significant in the hierarchical regression analysis. The regression model predicted 11.9% of the variance in knowledge (*F*_(4,401)_=13.51, *p*<0.001).

**Table 4.** Hierarchical regression analysis predicting knowledge.

| Model | B | SE | $\beta$ | *t* | $\Delta R^2$ |
| --- | --- | --- | --- | --- | --- |
| **Block 1 - Socio-demographics** ($F_{(3,402)}=17.32$; $p<0.001$) | | | | | 0.11 |
| Gender<sup>a</sup> | -0.82 | 0.25 | -0.16 | -3.30** |  |
| Education<sup>b</sup> | 0.71 | 0.24 | 0.14 | 2.94** |  |
| Marital status<sup>c</sup> | -1.35 | 0.35 | -0.19 | -3.85*** |  |
| **Block 2 – Observation checklist** ($F_{(1,404)}=3.6$; $p=0.058$) | | | | | 0.009 |
| Hand protection<sup>d</sup> | -1.08 | 0.78 | -0.07 | -1.39 |  |

Note: B=unstandardized regression coefficient; SE=Standard error; $\beta$ =standardized regression coefficient; <sup>a</sup>1=Male, 2=Female; <sup>b</sup>1= No formal education, 2=Primary level, 3=Secondary level; <sup>c</sup>1= Unmarried, 2=Married, 3=Divorced; <sup>d</sup>1=Yes, 2=No; $F_{(4,401)}=13.51$ , $p < 0.001$ , $R^2_{Adj}=0.11$ ; \*\* $p < 0.01$ , \*\*\* $p < 0.001$ .

Furthermore, the knowledge score was positively along with significantly correlated with the score of both attitudes (*r*=0.52) and practice (*r*=0.47).

### Attitude

The mean scores of attitudes were 12.3 (SD=1.7) out of 14 with an overall correct rate of 87.9 %. The distribution of each practices related question along with gender differences was presented in Table 5. The participants’ attitudes score was significantly higher among i) males vs. females (12.7±1.5 vs. 11.8±1.8, *p*<0.001), ii) participants with lower vs. upper ages (12.6±1.5 vs. 12.0±1.8, *p*=0.005), iii) day laborers vs. other occupation (12.8±1.5 vs. 11.4±1.8, *p*=0.013), iv) unmarried vs. divorced participants (12.7±1.3 vs. 10.8±2.1, *p*<0.001), and vi) participants those having monthly family income >15,000 BDT vs. <5,000 BDT (12.6±1.6 vs. 12.0±1.9, *p*=0.034) (see Table 3).

**Table 5.** Attitude and gender difference of participants (N=406)

| Characteristics | Total N=406 | | Male | | Female | | $\chi^2$ | df | *p*-value |
| --- | --- | --- | --- | --- | --- | --- | --- | --- | --- |
|  | *n* | (%) | *n* | (%) | *n* | (%) |  |  |  |
| Coronavirus is very serious |  |  |  |  |  |  |  |  |  |
| Disagree | 1 | (0.2) | 0 | (0.0) | 1 | (0.5) | 5.740 | 2 | 0.028* |
| Undecided | 20 | (4.9) | 6 | (2.8) | 14 | (7.4) |  |  |  |
| Agree | 385 | (94.8) | 210 | (97.2) | 175 | (92.1) |  |  |  |
| Coronavirus is preventable |  |  |  |  |  |  |  |  |  |
| Disagree | 1 | (0.2) | 1 | (0.5) | 0 | (0.0) | 14.766 | 2 | <0.001* |
| Undecided | 149 | (36.7) | 61 | (28.2) | 88 | (46.3) |  |  |  |
| Agree | 256 | (63.1) | 154 | (71.3) | 102 | (53.7) |  |  |  |
| Spatial distancing is mandatory to prevent COVID-19 |  |  |  |  |  |  |  |  |  |
| Disagree | 1 | (.2) | 0 | (.0) | 1 | (0.5) | 12.155 | 2 | 0.001* |
| Undecided | 94 | (23.2) | 36 | (16.7) | 58 | (30.5) |  |  |  |
| Agree | 311 | (76.6) | 180 | (83.3) | 131 | (68.9) |  |  |  |
| It is crucial to wear a face mask in crowded place |  |  |  |  |  |  |  |  |  |
| Disagree | 0 | (0.0) | 0 | (.0) | 0 | (0.0) | 0.001 | 1 | 0.103* |
| Undecided | 6 | (1.5) | 1 | (.5) | 5 | (2.6) |  |  |  |
| Agree | 400 | (98.5) | 215 | (99.5) | 185 | (97.4) |  |  |  |
| It is important to maintain home quarantine after getting infected COVID-19 |  |  |  |  |  |  |  |  |  |
| Disagree | 2 | (0.5) | 0 | (0.0) | 2 | (1.1) | 19.102 | 2 | <0.001* |
| Undecided | 209 | (51.5) | 91 | (42.1) | 118 | (62.1) |  |  |  |
| Agree | 195 | (48.0) | 125 | (57.9) | 70 | (36.8) |  |  |  |
| It can be treated at home |  |  |  |  |  |  |  |  |  |
| Disagree | 2 | (.5) | 1 | (.5) | 1 | (0.5) | 28.839 | 2 | <0.001* |
| Undecided | 208 | (51.2) | 84 | (38.9) | 124 | (65.3) |  |  |  |
| Agree | 196 | (48.3) | 131 | (60.6) | 65 | (34.2) |  |  |  |
| Safety measures can play an important role in COVID-19 prevention. |  |  |  |  |  |  |  |  |  |
| Disagree | 1 | (0.2) | 0 | (0.0) | 1 | (0.5) | 4.125 | 2 | 0.047* |
| Undecided | 3 | (0.7) | 0 | (0.0) | 3 | (1.6) |  |  |  |
| Agree | 402 | (99.0) | 216 | (100.0) | 186 | (97.9) |  |  |  |

Note: Fisher's exact test

**Table 6.** Hierarchical regression analysis predicting attitudes.

| Model | B | SE | $\beta$ | *t* | $\Delta R^2$ |
| --- | --- | --- | --- | --- | --- |
| **Block 1 - Socio-demographics** ($F_{(5,400)}=11.5$; $p<0.001$) | | | | | 0.12 |
| Gender<sup>a</sup> | -0.70 | 0.21 | -0.20 | -3.27** |  |
| Age<sup>b</sup> | -0.49 | 0.17 | -0.14 | -2.89** |  |
| Occupation<sup>c</sup> | -0.02 | 0.07 | -0.01 | -0.24 |  |
| Marital status<sup>d</sup> | -0.67 | 0.24 | -0.14 | -2.75** |  |
| Family income<sup>e</sup> | 0.28 | 0.14 | 0.10 | 2.03* |  |

Note: B=unstandardized regression coefficient; SE=Standard error; $\beta$ =standardized regression coefficient; <sup>a</sup>1=Male, 2=Female; <sup>b</sup>1=18-40 years, 2=>40 years; <sup>c</sup>1=House wife, 2=Workers, 3=Day laborer, 4=Shop keeping, 5=Rickshaw puller, 6=Others; <sup>d</sup>1= Unmarried, 2=Married, 3=Divorced; <sup>e</sup>1= <5,000 BDT, 2=6,000-15,000 BDT, 3= >15,000 BDT; $F_{(5,400)}=11.5$ ; $p<0.001$ , $R^2_{Adj}=0.11$ ; \* $p<0.05$ , \*\* $p<0.01$ .

Finally, factors that were statistically significant in the group difference analyses (t-tests and ANOVA) were included in a hierarchical regression analysis (see Table 6). Socio-demographic factors (i.e., gender, age, occupation, marital status, and family income) were included in Block 1. The more positive attitudes were positively associated with male (β=-0.20, *p*<0.01), lower age (β=-0.14, *p*<0.01), being unmarried (β=-0.14, *p*<0.01), and having higher income (β=0.10, *p*<.05). Consequently, occupation was not significant in the hierarchical regression analysis. The regression model predicted 11.6% of the variance in attitudes (*F*_(5,400)_=11.5, *p*<0.001).

Furthermore, the attitudes score was positively along with significantly correlated with the score of both knowledge (*r*=0.52) and practice (*r*=0.57).

### Practices

The mean scores of practices were 9.8 (SD=1.6) out of 12 with an overall correct rate of 81.7%. The distribution of each practices related question along with gender differences was presented in Table 7. The participants’ attitudes score was significantly higher among i) males vs. females (10.0±1.5 vs. 9.5±1.6, *p*=0.001), ii) participants with secondary level of education vs. no formal education (10.0±1.7 vs. 9.0±2.3, *p*<0.001), iii) day laborers vs. other occupation (10.5±1.5 vs. 9.3±1.8, *p*=0.026), iv) unmarried vs. divorced participants (9.9±1.7 vs. 8.5±2.0, *p*<0.001), v) participants living with nuclear vs. joint families (9.8±1.5 vs. 9.3±1.9, *p*=0.042), vi) participants those having monthly family income >15,000 BDT vs. <5,000 BDT (10.1±1.4 vs. 9.3±1.8, *p*<0.001), and v) participants those were observed with vs. without wearing face masks during the survey periods (9.8±1.4 vs. 9.5±2.1, *p*<0.001) (see Table 3).

**Table 7.** Practice and gender difference of participants (N=406)

| Characteristics | Total N=406 | | Male | | Female | | $\chi^2$ | df | p-value |
| --- | --- | --- | --- | --- | --- | --- | --- | --- | --- |
|  | n | (%) | n | (%) | n | (%) |  |  |  |
| Do you use tissues or hanker chips during coughing/sneezing? |  |  |  |  |  |  |  |  |  |
| Never | 1 | (0.2) | 1 | (0.5) | 0 | (0.0) | 3.176 | 2 | 0.155* |
| Occasionally | 26 | (6.4) | 10 | (4.6) | 16 | (8.4) |  |  |  |
| Always | 379 | (93.3) | 205 | (94.9) | 174 | (91.6) |  |  |  |
| Do you wear face mask when outing? |  |  |  |  |  |  |  |  |  |
| Never | 1 | (0.2) | 1 | (0.5) | 0 | (0.0) | 5.254 | 2 | 0.039* |
| Occasionally | 20 | (4.9) | 6 | (2.8) | 14 | (7.4) |  |  |  |
| Always | 385 | (94.8) | 209 | (96.8) | 176 | (92.6) |  |  |  |
| Do you avoid touching face and eyes? |  |  |  |  |  |  |  |  |  |
| Never | 1 | (0.2) | 0 | (0.0) | 1 | (0.5) | 3.851 | 2 | 0.084* |
| Occasionally | 33 | (8.1) | 13 | (6.0) | 20 | (10.5) |  |  |  |
| Always | 372 | (91.6) | 203 | (94.0) | 169 | (88.9) |  |  |  |
| Do you wash hands frequently using water and soaps? |  |  |  |  |  |  |  |  |  |
| Never | 0 | (0.0) | 0 | (0.0) | 0 | (0.0) | 1.458 | 1 | 0.252* |
| Occasionally | 102 | (25.1) | 49 | (22.7) | 53 | (27.9) |  |  |  |
| Always | 304 | (74.9) | 167 | (77.3) | 137 | (72.1) |  |  |  |
| Do you maintain spatial distancing? |  |  |  |  |  |  |  |  |  |
| Never | 11 | (2.7) | 4 | (1.9) | 7 | (3.7) | 1.302 | 2 | 0.523 |
| Occasionally | 53 | (13.1) | 29 | (13.4) | 24 | (12.6) |  |  |  |
| Always | 342 | (84.2) | 183 | (84.7) | 159 | (83.7) |  |  |  |
| Do you eat healthy food focusing on outbreak? |  |  |  |  |  |  |  |  |  |
| Never | 313 | (77.1) | 148 | (68.5) | 165 | (86.8) | 22.031 | 2 | <0.001 |
| Occasionally | 15 | (3.7) | 8 | (3.7) | 7 | (3.7) |  |  |  |
| Always | 78 | (19.2) | 60 | (27.8) | 18 | (9.5) |  |  |  |

Note: Fisher's exact test

**Table 8.** Hierarchical regression analysis predicting practice.

| Model | B | SE | $\beta$ | *t* | $\Delta R^2$ |
| --- | --- | --- | --- | --- | --- |
| **Block 1 - Socio-demographics** ($F_{(6,399)}=7.3$; $p<.001$) | | | | | 0.10 |
| Gender<sup>a</sup> | -0.28 | 0.20 | -0.09 | -1.40 |  |
| Education<sup>b</sup> | 0.30 | 0.15 | 0.09 | 1.95* |  |
| Occupation<sup>c</sup> | 0.00 | 0.06 | 0.00 | -0.01 |  |
| Marital status<sup>d</sup> | -0.68 | 0.23 | -0.16 | -2.94** |  |
| Family type<sup>e</sup> | -0.19 | 0.25 | -0.04 | -0.76 |  |
| Family income<sup>f</sup> | 0.41 | 0.13 | 0.16 | 3.17** |  |
| **Block 2 – Observation checklist** ($F_{(1,404)}=3.8$; $p=0.052$) | | | | | 0.009 |
| Using mask<sup>g</sup> | -0.25 | 0.19 | -0.06 | -1.3 |  |

Note: B=unstandardized regression coefficient; SE=Standard error; $\beta$ =standardized regression coefficient; <sup>a</sup>1=Male, 2=Female; <sup>b</sup>1=No formal education, 2=Primary level, 3=Secondary level; <sup>c</sup>1=House wife, 2=Workers, 3=Day laborer, 4=Shop keeping, 5=Rickshaw puller, 6=Others; <sup>d</sup>1=Unmarried, 2=Married, 3=Divorced; <sup>e</sup>1=Nuclear, 2= Joint; <sup>f</sup>1=<5,000 BDT, 2= 6,000-15,000 BDT, 3= >15,000 BDT; <sup>g</sup>1=Yes, 2=No; $F_{(7,398)}=6.5$ ; $p<0.001$ , $R^2_{Adj}=0.09$ ; \* $p<0.05$ , \*\*\* $p<0.001$ .

Finally, factors that were statistically significant in the group difference analyses (t-tests and ANOVA) were included in a hierarchical regression analysis (see Table 8). Socio-demographic factors (i.e., gender, education, occupation, marital status, family type, family income) were included in Block 1 Observation checklist (i.e., using mask) comprised Block 2. The more frequent practices were positively associated with a comparatively higher level of education (secondary level) (β=-0.09, *p*<0.05), being unmarried (β=-0.16, *p*<0.01), and having higher income (β=0.16, *p*<0.01). Consequently, gender, occupation, family type, and using masks were not significant in the hierarchical regression analysis. The regression model predicted 8.7% of the variance in attitudes (*F*_(7,398)_=6.5, *p*<0.001).

Furthermore, the practice score was positively along with significantly correlated with the score of both knowledge (*r*=0.47) and attitudes (*r*=0.57).

## Discussion

COVID-19 infections have continued to propagate exponentially with the wider community-level transmission in Bangladesh and are already ranked among the 20 countries with the highest transmission of COVID-19 (22). In such a catastrophic scenario, people with low incomes and the poorest, especially slum residents in Bangladesh, find themselves in the most vulnerable condition (23). Therefore, the present study aimed at investigating the level of knowledge, attitude, and practices (KAP) towards COVID-19 among slum dwellers at Dhaka city in Bangladesh. To the best of our knowledge, this is the first study in Bangladesh examining the KAP towards COVID-19 among slum dwellers. The study found that only 35.9% of slum dwellers were knowledgeable about COVID-19 mean scores of knowledge were 6.1 (SD=2.6), which implies that a majority of the Bangladeshi slum dwellers had inadequate knowledge about COVID-19 and this finding is not surprising due to the fact that slum dwellers are the marginalized people of the society with little education and low economic status (24,25).

Findings from this study showed that mass media (e.g., Television, Radio, etc.) was the main source of knowledge regarding the COVID-19. This is contrasting with recent KAP studies among Bangladeshi general people and found social media (e.g., Facebook) as the widely used source of knowledge regarding COVID-19 (15,19). This finding can be justified by the fact that the majority of the slum dwellers don’t have internet access as a result very few slum dwellers relying on social media such as Facebook to gather information regarding COVID-19. Therefore, it is suggested that a health education program incorporating mass media with a variety of advertisements can be influential in the dissemination of information on COVID-19.

A substantial number of sociodemographic and preventive factors significantly affect participant’s knowledge scores such as genders, education levels, marital status, and maintenance of hand protection. A previous KAP study in Bangladesh also reported that knowledge of COVID-19 significantly diverged across age, gender, education levels, residences, income groups, and marital status (16). Hierarchical regression analysis found participant’s favorable knowledge was positively associated with males, having a secondary level of education, and being unmarried. Male participants were more likely to have better knowledge about COVID-19. This finding is consistent with a previous KAP study among Bangladeshi internet users (26) but in contrast with studies conducted among Chinese and Saudi Arabian general people which reported females were more knowledgeable about COVID-19 (13,27). Moreover, higher education level was found to significantly associate with having accurate knowledge. Similar findings were also found in previous studies regarding COVID-19 in Bangladesh (15) and Iran (28). It is obvious due to the fact that educated persons are generally more aware of the potentiality of COVID-19 as a result they more knowledge. Interestingly, the present study found that participants who were unmarried having accurate knowledge compared to other groups which are consistent with previous KAP study among Bangladeshi people (16). A study conducted among Thai people found marital status, education, occupation, income level were significantly associated with accurate knowledge of COVID-19 (29), whereas a study among Chinese people found that female sex, age-group, marital status, education, and employment were significantly associated with knowledge (13).

The present study found that a large majority (87.9%) of participants showed a positive attitude toward COVID-19 with a mean attitude score of 12.3 (SD=1.7). Surprisingly, participants showed a positive attitude in terms of wearing a facemask, social distancing, and personal safety. A previous KAP study also found Bangladeshi people also showed a positive attitude towards wearing a facemask, frequent hand washing, and reporting to health authorities suspected cases of COVID-19 (20). But only 63.1% of slum dwellers knew COVID-19 is preventable which was contrasting to recent KAP study among Bangladeshi general people (90%) (20). Furthermore, Bangladeshi young adults perceived that COVID-19 will be successfully controlled, and confident about the government initiative to combat COVID-19 (19). A high level of positive attitude regarding COVID-19 is also found in studies conducted among people in Malaysia, China, and Saudi-Arabia (13,14,27). Positive attitudes towards COVID-19 can be interpreted by the Bangladesh government’s unprecedented precautionary steps including the lockdown, and suspension of all flights and the closing of all offices and educational institutions to safeguard citizens from COVID-19 (30). In the hierarchical regression analysis, sociodemographic variables associated with more positive attitudes regarding COVID-19 were being male, lower age, being unmarried, and having a higher income, overall recapitulating previous findings from China (13). In contrast, a similar study in Pakistan found participants’ attitudes were not affected by age, gender, experience, and job/occupation (31). Participants’ positive attitude was significantly correlated with the level of COVID-19 knowledge, similarly prior studies among citizens of Malaysia and Vietnam (14,32).

Although knowledge regarding COVID-19 was limited, the present study exhibits still a notable proportion (81.7%) of the slum dwellers in Bangladesh had good preventive practices to COVID-19 with a mean score of practices were 9.8 (SD=1.6). A large share of the participants (> 90%) reported using tissues or hanker chips during coughing/sneezing, wearing a face mask when outing, and avoid touching the face and eyes which is consistent with previous KAP study among Bangladeshi people (20). Less than 80% of participants stated that they always wash their hands using water and soaps which indicates a much indifference in practicing hand hygiene among the slum dwellers. This finding was remarkably similar to previous studies (27,33) except for a study in Thailand that found 54.8% did not regularly use soap during the washing of hands (29). More than 90% of participants reported to wearing masks when going outside the home which is similar to previous KAP studies regarding COVID-19 (26,28) but contradictory to a study conducted among Malaysian people which found 51.2% of participants reported wearing a face mask when going out in public (14). Results from hierarchical regression analysis found that more frequent practices were significantly associated with a higher level of education, being unmarried, having higher income, and Knowledge about COVID-19. Participants with a higher education showed more favorable preventive practices towards COVID-19. Similar findings were also found in previous studies (20,33). A recent KAP study in Bangladesh found participants’ age and sex are the factors associated with good practices towards COVID-19 (26). The study conducted among Malaysian citizens showed age, occupation, and income level, and knowledge regarding COVID-19 was correlated with good practices (14) whereas, sex, occupation, marital status, residence place, and COVID-19 knowledge was identified in a study in Chinese citizens (13). Therefore, tailoring the information provided by health officials and other media outlets on the disease needs to address the multifactorial nature of the drivers leading to reduced knowledge.

### Limitations

This study has several limitations which warrant consideration. This study followed a cross-sectional study design that cannot establish causal inferences. Therefore, a longitudinal study would overcome this limitation in understanding potential causal relationships. Moreover, the modestly sized sample is only representative of the selected slums in Dhaka City and might not be representative of other areas or other countries. Therefore, studies utilizing bigger samples from more representative populations are needed.

## Conclusions

In essence, findings from this study revealed that the majority of the slum dwellers in Bangladesh have limited knowledge of the COVID-19. Interestingly, they exhibit positive attitudes and favorable practices towards COVID-19. But these are not satisfactory they are still in a life-threatening condition due to their cramped housing condition where preventive practices such as personal hygiene and maintaining physical distancing are unrealistically impossible. Furthermore, lack of knowledge regarding COVID-19 is another matter of concern. Therefore, immediate implementation of and culturally sensitive health education together with better housing and providing adequate facilities favorable for precautions against COVID-19 is desperately needed to help people encourage an even more positive mindset and maintain appropriate preventive practices and combat against this pandemic.

## Data Availability

All data are fully available without any restriction upon reasonable request.

## Acknowledgments

The authors would like to thank all of the participants who consented willingly and enrolled in the study voluntarily.

## Authors’ contribution

**Md. Saiful Islam:** Conceptualization, Methodology, Investigation, Data curation, Formal analysis, Writing - original draft, Editing, Validation., **Md. Galib Ishraq Emran:** Conceptualization, Investigation, Writing - original draft, Validation., **Md. Estiar Rahman:** Writing - original draft, Editing, Validation., **Rajon Banik:** Writing - original draft, Editing, Validation., **Md. Tajuddin Sikder:** Editing, Validation., **Lee Smith:** Editing, Validation., **Sahadat Hossain:** Supervision, Conceptualization, Investigation, Editing, Validation.

## Ethical approval

All procedures of the present study were carried out in accordance with the principle for human investigations (i.e., Helsinki Declaration). Formal ethics approval was granted by the ethical review board of the Faculty of Biological Sciences, Jahangirnagar University, Savar, Dhaka-1342, Bangladesh (Ref. No: BBEC, JU/M 2020/COVID-19/(8)5).

## Informed consent

Each participant consented willingly during the interview period.

## Data availability statement

All data are fully available without any restriction upon reasonable request.

